# Clinical outcomes of HIV treatment clients enrolled in six-month dispensing after less than 6 months on treatment in Zambia: A target trial emulation

**DOI:** 10.64898/2026.07.30.26358576

**Authors:** Elizabeth Kachingwe, Matthew P. Fox, Idah Mokhele, Khumbo Shumba, Vinolia Ntjikelane, Suilanji Sivile, Aniset Kamanga, Prudence Haimbe, Sydney Rosen, Sophie Pascoe, Amy Huber

## Abstract

**Background:** Six-month multi-month dispensing (6MMD) of antiretroviral therapy (ART) reduces clinic visit frequency and is associated with improved retention in care. During the COVID-19 pandemic, Zambia offered 6MMD to clients ≥3 months after ART initiation, rather than the ≥6 months standard requirement. We estimated the effect of early (<6 months on ART) versus standard (6-12 months) 6MMD enrolment on rate of treatment interruption.

**Methods:** We emulated a target trial using routinely collected electronic medical records from 12 public health facilities in Zambia. Eligible clients were ≥15 years, initiated ART 01/20-08/22, were WHO stage 1 or 2 at ART initiation, and had ≥21 months of potential follow-up. Treatment interruption was defined as missing a scheduled clinic or pharmacy visit by >28 days. We applied a clone-censor-weight approach to reduce immortal time bias. Clones were censored when observed dispensing deviated from their assigned strategy. Inverse probability of censoring weights (IPCW) accounted for informative censoring, while inverse probability of treatment weights (IPTW) balanced measured baseline confounders between strategies. We used weighted pooled logistic regression of person-month data to estimate the odds of treatment interruption between early and standard 6MMD enrollers, including follow-up month to model the monthly baseline risk.

**Results:** 6,142 ART clients met inclusion criteria. 741 (12.1%) were early 6MMD enrollers, 1,590 (26.1%) standard 6MMD enrollers, and 3,811 (62.0%) eligible clients who never enrolled in 6MMD. During follow-up, 268 treatment interruptions occurred. In the primary analysis, early 6MMD was associated with lower odds of treatment interruption than standard 6MMD OR 0.701 (95% CI 0.51– 0.97). The predicted cumulative probability of treatment interruption at 18 months was 6.5% under the early 6MMD strategy and 9.1% under the standard strategy (risk difference: −2.6 percentage points).

**Conclusions:** Enrolment in 6MMD at 3-6 months after ART initiation was associated with lower odds of treatment interruption than standard enrolment at 6-12 months, with a predicted absolute risk difference of −2.6 percentage points at 18 months. We found no evidence that earlier access to 6MMD increases the risk of treatment interruption.

## INTRODUCTION

Multi-month dispensing (MMD) of antiretroviral therapy (ART) has been widely implemented to enhance HIV treatment adherence, improve retention in care, and optimize healthcare resource utilization. The World Health Organization (WHO) first recommended 3- to 6-month dispensing intervals for clinically stable individuals in 2016, later labelling 6-month multi-month dispensing (6MMD) the preferred strategy in 2021[1,2]. Eligibility for 6MMD has typically been restricted to adults considered stable or established on ART, typically defined as having been on ART ≥1 year. Empirical evidence suggests 6MMD is associated with improved viral load suppression, reduced loss to follow-up, and increased patient satisfaction, while also alleviating healthcare facility congestion and reducing provider workload. Additionally, 6MMD has been linked to cost savings for both patients and health systems[3,4].

In line with international recommendations, in 2018 the Zambian Ministry of Health (MOH) implemented a nationwide scale-up of MMD models[5,6]. Revised ART guidelines prioritized expanding access to multi-month dispensing and decentralizing ART delivery through community health workers, local pharmacies, and mobile clinics[7]. In March 2020, anticipating pandemic-related disruptions to healthcare access, Zambia and several other low- and middle-income countries (LMICs)[8–11] adopted early six-month ART refills to minimize clinic visits and reduce the risk of COVID-19 exposure for people living with HIV[12]. Zambia started permitting six-month dispensing after 3 months on ART in March 2020[13].

Empirical studies examining the impact of MMD on retention in sub-Saharan Africa have yielded mixed findings. While some research indicates that MMD offered after 6 months on ART improves adherence and retention while reducing clinic congestion, others found that extended refill intervals had no impact on retention[14]. Very limited data exist on the effects of earlier access to MMD (i.e. <6 months on ART) compared to the standard enrolment threshold of ≥6 months. Newly initiated ART clients, particularly those within the first six months of treatment, are generally at heightened risk for early disengagement, suboptimal adherence, and loss to follow-up compared to those who have been on ART for a longer duration[15]. The role of varying dispensing intervals to this risk is unknown.

Comparing early enrollers, those who initiate 6MMD soon after starting ART, to individuals who enrol in 6MMD after longer ART exposure can provide critical insights to guide potential expansion of eligibility criteria for countries in similar settings. Prior studies suggest that newly initiated clients prefer fewer clinic visits soon after ART initiation, indicating that early access to 6MMD may enhance satisfaction and engagement in care[16,17]. This study estimates the effect on treatment interruptions of early enrolment into MMD compared to standard enrolment, using a target trial emulation framework.

## METHODS

### Target trial specification

Target trial emulation specifies the hypothetical randomised trial that an observational study is designed to approximate, including eligibility criteria, treatment strategies, time zero, follow-up, and the causal contrast of interest. This framework helps reduce biases common in observational analyses, including immortal time bias. For this study, our target trial compares early 6MMD enrolment after 3 months on ART with standard enrolment after 6 months on ART, estimating the effect on treatment interruption. To avoid excluding treatment interruptions occurring between 3 and 6 months among those eligible for the standard enrolment strategy, we used a clone-censor-weight approach that includes all eligible clients from time zero. Table 1 outlines the design of the target trial and the corresponding specifications for its emulation using observational data.

**Table 1:** Specification of a target trial to compare effect on the rate of interruption in treatment of early enrolment in 6MMD with standard enrolment.

| Protocol component | Randomized trial | Emulated trial |
| --- | --- | --- |
| Eligibility criteria | Clients $\geq 15$ years old who started ART between 1 January 2020 and August 2022 at one of 12 public health facilities in Zambia, 3 to 6 months on ART, were not pregnant or WHO stage 3 or 4 at ART initiation, and no transfer-in history | Same as trial but had to have at least 18 months of potential follow-up before dataset closure |
| Dispensing strategies | Initiate 6MMD early (between 3 - <6 months on ART) or late (wait and initiate 6MMD 6 - <12 months on ART) | Same as trial |
| Assignment procedures | Participants randomized to early or standard enrolment into 6MMD at 3 months visit | Participants cloned and treatments assigned to the two clones of each participant between 3 to 6 months on ART |
| Time zero | 3 months post ART initiation | Same as trial |
| Follow-up period | Follow up until earliest of loss to follow-up, transferring out, death, or end of observation period 21 months after ART initiation | Same as trial except that participant clones are censored when they deviate from assigned dispensing strategies |
| Outcome of interest | Rate of treatment interruption up to 21 months after ART initiation | Same as trial |
| Causal contrasts of interest | Per-protocol effect (i.e., effect of adhering to the dispensing strategy assigned) | Same as trial |
| Analysis plan | Per-protocol analysis using survival analysis to assess the rate of treatment interruption where observations are censored when they deviate from the assigned treatment strategy and weighted to account for the censoring, | Per-protocol analysis using clone-censor-weight approach to create two copies of each individual at 3 months and assigning one to the early treatment strategy and the second to the standard treatment strategy. Each individual contributes person-time from time zero and clones are censored when they deviate from the defined strategy. We used inverse probability of censoring weights to address informative censoring. |

### Data sources

We utilized data from Zambia’s electronic medical record system (EMR), SmartCare[18], obtained from 12 public healthcare clinics located in Central and Lusaka Provinces. The dataset included HIV diagnosis information, baseline laboratory results, pharmacy dispensing records, follow-up visit data, treatment history, and data related to service delivery models for clients initiating ART.

### Eligibility criteria

In our emulated trial we included non-pregnant clients ≥15 years who initiated ART between January 2020 and August 2022, were classified in WHO stage 1 or 2 at ART initiation, completed ≥3 months on ART, and had ≥21 months of potential follow-up before dataset closure on 31 May 2024.

### Time zero and follow-up

We defined the start of follow-up (time zero) as three months after ART initiation. Follow-up continued until the earliest of 21 months post-initiation, last recorded visit (for those who interrupted treatment, were lost to follow-up, or died), or database closure on 31 May 2024.

### Treatment strategies

We compared two strategies for 6MMD enrolment timing: early enrolment, defined as initiating 6MMD between 3 and 6 months after ART initiation, versus standard enrolment, defined as initiating between 6 and 12 months after ART initiation. Clients were classified as having enrolled in 6MMD if their records indicated a six-month dispensing interval, documented enrolment in a differentiated service delivery model incorporating six-month refills, or a dispensed quantity of antiretrovirals corresponding to six months of treatment. These criteria were applied to account for variation in how dispensing information was recorded across facilities. The first instance of 6MMD was considered the 6MMD enrolment date for each client.

### Outcome

The primary outcome was treatment interruption, defined as missing a scheduled clinic or pharmacy visit by more than 28 days, consistent with Zambian ART guidelines. Treatment interruption was treated as a terminal event, ending an individual client’s follow-up at the point it occurred.

### Target trial emulation and statistical analysis

Directly comparing clients who enrolled in 6MMD early versus later using routine program data can introduce bias. Clients who enrol later must have remained in care long enough to reach that later enrolment window, creating immortal time bias. In addition, clients who enrol early may differ systematically from those who enrol later, resulting in confounding.

To address these challenges, we used a clone–censor–weight approach to emulate a target trial. At time zero, each eligible client was cloned and assigned to both enrolment strategies, emulating baseline randomisation. Clones were censored when observed care deviated from their assigned strategy. For example, a client initiating 6MMD during the standard enrolment window would be censored in the clone assigned to the early enrolment strategy at that time. Supplementary Figure 1 illustrates this approach using seven hypothetical clients.

Because artificial censoring may be informative (i.e., clients who deviate from their assigned strategy and are therefore censored may differ systematically from those who remain on-strategy, meaning censoring is not random with respect to the outcome), we estimated stabilised inverse probability of censoring weights (IPCW) using pooled logistic regression models that included follow-up time, treatment strategy, and baseline client and facility characteristics. Propensity scores were estimated using logistic regression, with age at ART initiation, sex, facility ART cohort size, and urban or rural facility location as covariates. These propensity scores were used to derive stabilised inverse probability of treatment weights (IPTW) to balance measured baseline characteristics between the early and standard enrolment groups. Covariate balance after weighting was assessed using standardised mean differences; values < 0.10 indicated adequate balance. Final weights were the product of the IPCW and IPTW. To limit the influence of extreme values, weights were trimmed at the 99th percentile.

We fitted two weighted pooled logistic regression models to person-month data to estimate the association between enrolment strategy and treatment interruption: one weighted by IPCW alone, and one by the combined IPTW × IPCW weight. Both models included treatment strategy and follow-up month, with robust standard errors to account for correlation between clones. From the fully weighted model, we estimated 18-month (from time zero) cumulative incidence by strategy and calculated the corresponding risk difference.

As a sensitivity analysis, we fitted weighted Poisson regression models using the same weighting approach and compared results with those from the pooled logistic regression models. Full model specifications are provided in the Supplementary Methods (Supplementary Table 3).

### Ethical considerations

Permission to access anonymized routine data from the Smart Care EMR system was obtained from the Zamiba Ministry of Health (MOH). The study received ethical approval from three institutional review boards: the Zambia Research Committee (2019-Sep-030) Boston University Institutional Review Board (IRB Number: H-38822), and the University of the Witwatersrand Human Research Ethics Committee (Ref: M240911). Only anonymised data were accessed by the research team.

## RESULTS

### Study cohort

Of the 62,843 registered ART clients, 6,142 (9.8%) met our eligibility criteria and were included in the analysis. Exclusions were due to ART initiation after 2022 (n=15,361; 24.4%) or before 2020 (n=14,032; 22.3%), WHO stage 3/4 or missing WHO stage information (n=9,111; 14.5%), missing ART start dates (n=5,737; 9.1%), and other reasons listed in Figure 2. Among eligible clients, 1,590 (25.9%) initiated standard 6MMD, 741 (12.1%) initiated early 6MMD, and 3,811 (62.0%) were eligible but did not initiate 6MMD during the study period.

**Figure 1.**
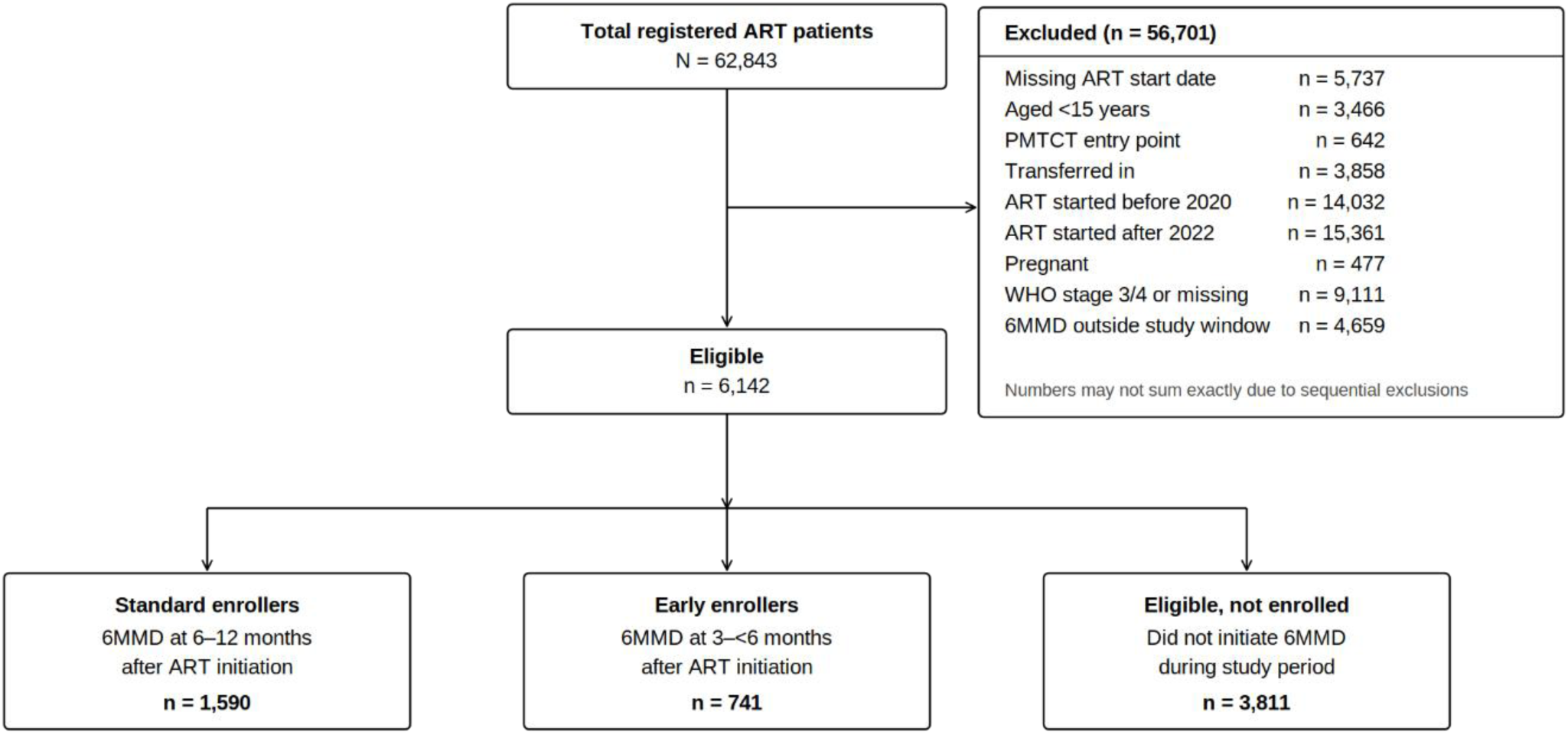
Selection of clients ≥15 years old who initiated treatment between 1 January 2020 to 31 December 2022

The median (IQR) age of the cohort was 32 years (26–39). Standard and early enrollers were older, with a median age of 34 years (27–41 and 28–42, respectively), than were clients who never enrolled (31, 26–38). Among clients aged 15–24 years, 70.3% never enrolled in 6MMD, while 20.8% were standard enrollers and 8.9% were early enrollers. Among clients aged 45–54 years, 51.8% never enrolled, 32.1% were standard enrollers and 16.1% were early enrollers. Among those aged 55 years and older, 45.7% never enrolled in 6MMD, 37.0% were standard enrollers and 17.4% were early enrollers. Table 2 presents characteristics of the cohort and study facilities. Females comprised approximately two thirds of the cohort, consistent with the overall Zambian ART population, but were less likely than males to enrol in 6MMD during follow-up.

**Table 2.** Characteristics of eligible clients at time zero.

| Characteristic | Standard enrollers<br>(enrolled at 6-12 months) | Early enrollers<br>(enrolled at 3-6 months) | Eligible, never enrolled in 6MMD | Total |
| --- | --- | --- | --- | --- |
| N (row %) | 1,590 (26.1) | 741 (12.1) | 3,811 (62.0) | 6,142 |
| Median age (IQR) | 34 (27-41) | 34 (28-42) | 31 (26-38) | 32 (26-39) |
| Age (n, row %) |  |  |  |  |
| 15-24.9 years | 244 (20.8%) | 104 (8.9%) | 825 (70.3%) | 1,173 |
| 25-44.9 years | 1,109 (26.1%) | 526 (12.4%) | 2,613 (61.5%) | 4,248 |
| 45-54.9 years | 173 (32.1%) | 87 (16.1%) | 279 (51.8%) | 539 |
| 55+ years | 51 (37.0%) | 24 (17.4%) | 63 (45.7%) | 138 |
| Sex (n, row %) |  |  |  |  |
| Female | 932 (22.8%) | 475 (11.6%) | 2,684 (65.6%) | 4,091 |
| Male | 645 (32.1%) | 266 (13.3%) | 1,096 (54.6%) | 2,007 |
| Months on ART at 6MMD enrolment (median, IQR) | 9.1 (8-11) | 4.3 (3.5-5.3) |  | 8.2 (3.9-10.0) |
| ART initiation year (n, row %) |  |  |  |  |
| 2020 | 888 (27.7%) | 413 (12.9%) | 1,909 (59.5%) | 3,210 |
| 2021 | 613 (26.6%) | 274 (11.9%) | 1,414 (61.5%) | 2,301 |
| 2022 | 89 (14.1%) | 54 (8.6%) | 488 (77.3%) | 631 |
| Facility ART patient volume (n, row %) |  |  |  |  |
| 2000-4000 | 379 (30.1%) | 317 (25.1%) | 565 (44.8%) | 1,261 |
| >4000 | 1,211 (24.8%) | 424 (8.7%) | 3,246 (66.5%) | 4,881 |
| Facility setting (n, row %) |  |  |  |  |
| Urban | 1,463 (26.8%) | 670 (12.3%) | 3,328 (60.9%) | 5,461 |
| Rural | 127 (18.6%) | 71 (10.4%) | 483 (70.9%) | 681 |

\* ART, Anti-retroviral therapy; 6MMD, six months dispensing; IQR, interquartile range

Among all clients enrolled in 6MMD, the median time on ART at enrolment was 8.2 months (IQR: 3.9–10.0) overall. Early enrollers entered 6MMD at a median of 4.3 months (IQR: 3.5–5.3) after ART initiation, while standard enrollers entered at a median of 9.1 months (IQR: 8–11).

### Weighting diagnostics

Propensity scores were estimated before cloning to predict early versus standard 6MMD enrolment using age category, sex, facility type, and urban/rural location. Propensity score distributions showed substantial overlap between groups, supporting the positivity assumption. After trimming at the 99th percentile, stabilised IPTW weights were stable, with means close to 1.0 in both the early (0.992, SD 0.287) and standard (1.003, SD 0.153) arms, indicating no evidence of extreme weights.

IPCW weights were also stable, early: mean 0.995 (SD 0.020); standard: mean 1.002 (SD 0.008)). The narrow distribution of IPCW suggested limited informative censoring after conditioning on treatment strategy and baseline covariates. Combined IPTW × IPCW weights were similarly stable, with means close to 1.0 in both groups and no indication of positivity violations or model misspecification.

Before weighting, the greatest imbalance was observed for facility type (SMD = −0.410), followed by sex (SMD = −0.103); age category (SMD = 0.036) and urban/rural location (SMD = 0.056) were reasonably balanced. After IPTW, all measured baseline covariates were well balanced, with weighted SMDs below 0.01: facility type (−0.003), sex (−0.007), age category (0.005), and urban/rural location (0.001). These findings indicate that the weighting approach achieved good balance in measured baseline characteristics between the two enrolment strategies.

### Effect of early versus standard 6MMD on treatment interruption

Table 3 presents the results from the two weighted pooled logistic regression models used to estimate the effect on treatment interruption of early vs later 6MMD enrolment.

**Table 3.** Weighted treatment interruption estimates by model and treatment strategy.

| Model | Weighting | Effect estimate | 95% CI* |
| --- | --- | --- | --- |
| Pooled logistic regression primary model |  |  |  |
| Pooled logistic regression Model 1 | IPCW only | 0.614 | 0.450 – 0.837 |
| Pooled logistic regression Model 2 | IPTW + IPCW | 0.701 | 0.508 – 0.968 |
| Cumulative treatment interruption incidence at 18 months (PLR Model 2, IPTW + IPCW) |  |  |  |
| Standard 6MMD | IPTW + IPCW | 9.1 cumulative treatment interruption probability |  |
| Early 6MMD | IPTW + IPCW | 6.5 cumulative treatment interruption probability |  |
| Risk difference | IPTW + IPCW | –2.6 percentage points (early vs standard) |  |
| Reference - Poisson regression (cross-check) |  |  |  |
| Poisson Model 1 | IPCW only | 0.617 (IRR) | 0.453 – 0.842 |
| Poisson Model 2 | IPTW + IPCW | 0.703 (IRR) | 0.509 – 0.970 |

\*CIs are from the robust sandwich estimator. OR, odds ratio approximating hazard ratio under rare-event assumption; IRR, incidence rate ratio (Poisson cross-check); IPCW, inverse probability of censoring weighting; IPTW, inverse probability of treatment weighting; treatment interruption; 6MMD, six-monthly multi-month dispensing. PLR results from person-month data using stsplit at(0(1)21); Poisson results from same dataset with exposure offset.

In the IPCW-only model (Model 1), early 6MMD enrolment was associated with lower odds of treatment interruption than standard 6MMD enrolment (OR 0.614, 95% CI 0.450–0.837). After additional adjustment for measured baseline confounding using IPTW (Model 2 - IPCW + IPTW), the association was modestly attenuated but remained protective (0.701, 0.508–0.968).

This change in effect suggests that measured baseline differences between the groups contributed modestly to the unadjusted association; however, the direction and magnitude of the association remained broadly consistent across models, providing evidence that early enrolment into 6MMD was not associated with an increased risk of treatment interruption.

In sensitivity analysis, weighted Poisson regression produced near identical estimates (Model 1 IRR 0.617; 95% CI 0.453–0.842; Model 2 IRR 0.703, 95% CI 0.509–0.970), supporting the robustness of the findings to the modelling approach.

## DISCUSSION

In this analysis, we used routinely collected electronic medical records data from 12 health facilities in Zambia to emulate a target trial comparing early (3-<6 months after ART initiation) versus standard (6-12 months) enrolment into six-month multi-month dispensing (6MMD) of antiretroviral therapy, using a clone-censor-weight approach. Overall, our results suggest that earlier access to 6MMD does not increase the risk of treatment interruption and may offer a modest protective benefit among clients who initiated ART with WHO stage 1 or 2 disease and had completed at least three months of ART. In the primary analysis using pooled logistic regression (PLR) with inverse probability of censoring weighting only (Model 1), early 6MMD enrolment was associated with a 39% lower odds of treatment interruption compared with standard enrolment. After additional adjustment for baseline confounding through the combined inverse probability of treatment and censoring weights (Model 2), the effect estimate was modestly attenuated but remained consistent with a lower odds of treatment interruption, with early enrollers experiencing a 30% lower odds of treatment interruption than standard enrollers

On the absolute scale, the predicted cumulative probability of treatment interruption at 18 months was 9.1% for the standard 6MMD strategy and 6.5% for the early 6MMD strategy, corresponding to a risk difference of 2.6 percentage points in favour of early enrolment The sensitivity analysis demonstrated that the findings were robust, with the Poisson models yielding results consistent with those from the pooled logistic regression models. The change in effect estimate following adjustment for baseline confounding suggests that some of the apparent benefit observed in the IPCW-only analysis (Model 1) was attributable to differences in patient characteristics between treatment groups. Prior to weighting, early enrollers were more likely to attend smaller facilities and were slightly more likely to be female, characteristics that may independently influence retention in care. Weighting successfully balanced all measured baseline covariates, however, with standardised mean differences below the conventional threshold of 0.10. The persistence of a protective association after adjustment provides reassurance that the observed findings were not solely driven by measured differences between groups. Residual confounding by unmeasured characteristics cannot be excluded, however. For example, clinicians’ expectations of a client’s future adherence behaviour may affect enrolment. If clinicians are more likely to offer early 6MMD to those they judge to be “ready”, and assuming that their judgment is generally correct, then the early 6MMD group may include a larger proportion of clients with a lower underlying risk of interruption, which could partly explain the observed protective association.

Our findings are consistent with previous evidence from sub-Saharan Africa demonstrating that multi-month dispensing does not adversely affect treatment outcomes. Studies conducted in Malawi, Uganda, and Zambia have shown that 3- and 6-month dispensing models can maintain or improve retention in care and viral suppression compared with more frequent refill schedules[19–21]. Similarly, prior analyses from Zambia found no increase in loss to follow-up among clients initiated on multi-month dispensing soon after treatment initiation[22]. The INTERVAL cluster-randomized trial demonstrated that 6-month dispensing was non-inferior to 3-month dispensing among stable ART clients in Malawi and Zambia, though it did not enrol those on ART for less than 6 months[23].

Our study extends the existing evidence base by directly comparing two different timing strategies for six-month dispensing using a causal inference framework designed to emulate a randomised trial. The use of PLR within the clone-censor-weight framework allowed us to estimate both relative (odds ratio approximating hazard ratio) and absolute (cumulative incidence, risk difference) measures of effect, providing a more complete picture of the clinical and programmatic relevance of early 6MMD enrolment. The use of routinely collected programme data, moreover, allowed us to evaluate intervention effectiveness under real-world implementation conditions, where patient populations, clinical decision-making, and service delivery more closely reflect routine practice [14]. As a result, our findings provide evidence that earlier access to 6MMD is safe and may offer additional benefits beyond those previously reported.

Several mechanisms may explain the lower rate of treatment interruption observed among early enrollers. Receiving a six-month ART refill within the first three to six months of treatment initiation substantially reduces the frequency of clinic visits during a period when patients are still adapting to lifelong care. Fewer clinic visits may reduce barriers such as transportation costs, time away from work, childcare responsibilities, and other competing demands that can contribute to disengagement from care. Early enrolment into 6MMD may also signal providers’ trust in patients’ ability to manage their treatment, potentially strengthening patient engagement and satisfaction. Furthermore, reducing the number of required clinic encounters during the early treatment period may decrease opportunities for missed visits and subsequent treatment interruption[24].

The period between three and six months after ART initiation has traditionally been viewed as a time of elevated risk for disengagement, during which frequent clinical monitoring was considered necessary[15,25]. Although our findings do not directly evaluate the role of clinical monitoring during this period, they suggest that earlier access to 6MMD does not increase the risk of treatment interruption within the context of a mature ART programme such as Zambia’s, where differentiated service delivery models are well established, drug supply chains are reliable. In this setting, providing extended ART refills earlier in the treatment course appears to be safe and may be beneficial for selected clients who initiated ART with WHO stage 1 or 2 disease and completed at least three months of ART.

This study has several important strengths. First, we used a large routine clinical dataset drawn from 12 public health facilities, providing evidence from a real-world implementation setting. Second, the target trial emulation framework allowed us to explicitly define eligibility criteria, treatment strategies, follow-up, and outcomes, reducing common biases associated with observational analyses. Third, the clone-censor-weight approach addressed immortal time bias while inverse probability weighting accounted for both informative censoring and measured baseline confounding. Fourth, the use of PLR as the primary outcome model enabled estimation of both relative and absolute measures of effect at a pre-specified time point, strengthening the clinical interpretability of the findings. Finally, extensive weight diagnostics demonstrated adequate overlap between treatment groups, good covariate balance after weighting, and no evidence of substantial positivity violations, increasing confidence in the validity of the findings.

Alongside these strengths, our study also had several limitations. First, our analysis relied on routinely collected SmartCare electronic medical record data, which were not collected primarily for research purposes and may therefore be subject to incomplete or inaccurate documentation of clinical visits, dispensing information, and patient characteristics. Data quality and completeness may have varied across facilities and may have been further affected by operational disruptions during the COVID-19 pandemic, including changes in service delivery and documentation practices. Consequently, misclassification of 6MMD enrolment timing, outcomes, and covariates may have occurred. Second, although the target trial emulation framework and weighting approach reduced several important sources of bias, residual confounding due to unmeasured factors such as baseline CD4 count, treatment support, socioeconomic status, or individual motivation cannot be excluded.

Third, treatment interruption may have been misclassified because the SmartCare database could not fully capture unlinked inter-facility transfers (“silent transfers”) outside the 12 study sites. Consequently, some patients who re-engaged in HIV care elsewhere may have been incorrectly classified as experiencing an interruption in treatment. If such silent transfers occurred similarly across both treatment strategies, the estimated effect would likely have been biased toward the null. Conversely, if the longer medication supply provided through early 6MMD enabled clients to maintain treatment continuity during periods of geographic mobility, this would represent a genuine benefit of the intervention rather than a source of bias.

Fourth, enrolment into early 6MMD during the COVID-19 pandemic may have been influenced by clinical judgement, potentially resulting in selection of patients perceived to be more stable and adherent, thereby overestimating the benefits of early enrolment. Although weighting balanced measured covariates, clinicians may have preferentially enrolled patients perceived to be highly adherent, socially stable, or otherwise at lower risk of disengagement. Such unmeasured factors could have biased the estimated protective effect of early 6MMD away from the null. Fifth, the exclusion of patients with missing WHO staging information may have affected the representativeness of the study population and introduced selection bias if excluded patients differed systematically from those included. Sixth, the study was restricted to clients who initiated ART with WHO stage 1 or 2 disease and completed at least three months of ART, attending 12 sentinel sites in Central and Lusaka provinces and may not be generalisable to patients with advanced HIV disease or to facilities in other regions of Zambia. Finally, confidence intervals were derived from the analytic robust sandwich estimator; bootstrap confidence intervals were not computed, and the uncertainty around the magnitude of the effect should be interpreted accordingly.

Despite these limitations, from a programmatic perspective, our findings suggest that requiring six months of ART exposure before eligibility for 6MMD may not be necessary for patients who do well in their first three months. Expanding eligibility criteria to allow enrolment after three months of ART could reduce clinic burden, improve patient convenience, and support the goals of differentiated service delivery without compromising treatment continuity. Such an approach may also strengthen the resilience of HIV treatment programmes by reducing demands on both patients and health systems.

## Data Availability

The data that support the findings of this study are available from the Zambia Ministry of Health subject to the appropriate approvals and data-sharing agreements. The data are not publicly available because they contain confidential patient information.

https://smartcarezambia.io/

## Competing interests

The authors declare that they have no competing interests. PM is an employee of the government agency that has authority over the study sites.

## Financial disclosure

This study was supported by funding from the Gates Foundation through grant INV-037138 to the Wits Health Consortium and grant INV-031690 to Boston University. The funders had no role in the study design, data collection, analysis, interpretation of the results, or preparation of the manuscript.

## Authors’ contributions

*EK, ANH, IM, KS, and MPF conceptualised the study. AK, PH, ANH and SP facilitated remote data access. EK, MPF, ANH, KS, and IM developed the target trial emulation framework. EK conducted the data analysis with guidance from MPF and ANH. EK drafted the initial manuscript. MPF, SR, SP, ANH, KS, AK, VN, and PH critically reviewed and revised the manuscript. All authors contributed to the interpretation of the findings, reviewed the manuscript, and approved the final version*.

## Acknowledgements

The authors gratefully acknowledge the Zambia Ministry of Health for granting access to the data extracts and for providing ongoing technical support throughout this work. We also sincerely thank the Clinton Health Access Initiative (CHAI) Zambia team and IHM for facilitating access to the data and providing technical support with data management, interpretation of the analytical outputs, and implementation insights. We further acknowledge the healthcare workers, facility management, and patients at the participating primary healthcare facilities in Lusaka and Central Provinces for their support, collaboration, and contribution to this work.

---

**Supplementary table 3.** Target Trial Emulation full statistical analysis specifications to accompany the simplified methods section in the main manuscript. Sections follow the order of the main methods and expand on each analytic step in detail.

| Section | Topic | Detail |
| --- | --- | --- |
| S1 | Time zero and Follow-up | Time zero was defined as the time at which a client became eligible for the emulated trial: 3 months after ART initiation. This represents the start of follow-up in the target trial emulation, the point at which hypothetical randomisation occurs in the cloning approach. Follow-up continued through the earliest of 21 months post-ART initiation, last visit date (for those who interrupted treatment, were lost to follow-up, or died), or database closure on 31 May 2024.<br>Clients who were eligible for 6MMD but did not enrol at any point during the study period were censored at the end of the treatment assignment window corresponding to the strategy arm they were assigned in the cloning approach. |
| S2 | Treatment Strategies and 6MMD Classification | The exposure of interest was early enrolment into 6MMD compared to standard enrolment. **Early enrolment** was defined as initiating 6MMD between 3 and 6 months after ART initiation (0–3 months after time zero). **Standard enrolment** was defined as waiting to initiate 6MMD between 6 and 12 months after ART initiation (3–9 months after time zero).<br>Clients were classified as enrolled into 6MMD if their clinical records indicated a six-month dispensing interval, documented enrolment in a differentiated service delivery (DSD) model supporting six-month refills, or a quantity of antiretroviral (ARV) drugs dispensed corresponding to approximately six months of treatment. These criteria were applied to capture all clients receiving six-month ART refills, accounting for variation in how dispensing information was recorded across facilities. The first instance of 6MMD was used for each client. |
| S3 | Outcome | The primary outcome was interruption in treatment, defined as missing a scheduled clinic or pharmacy visit by more than 28 days, based on Zambian ART guidelines. Treatment interruption was treated as a terminal event: follow-up ended at the point it occurred and clients did not re-enter follow-up after interruption. |
| S4 | Rationale for the Clone-Censor-Weight Approach | Directly comparing clients who enrolled in 6MMD early versus later using routine programme data introduces two sources of bias. First, at time zero it is not yet known which treatment strategy a given client will follow. If only clients who initiated 6MMD within a given window were assigned to that strategy group, those assigned to the standard enrolment group (6–12 months after ART initiation) would have had to survive, remain in care, and be outcome-free for 3 full months after time zero before becoming eligible generating immortal person-time that is incorrectly attributed to that group. For example, an individual who initiates 6MMD at 2 months after time zero could legitimately have been assigned to the early enrolment strategy (0–3 months) but could also have been in the standard enrolment group for the first 2 months. Assigning them only to the early group ignores this ambiguity and introduces bias.<br>Second, a client who never initiates 6MMD could plausibly have been allocated to either strategy. Excluding non-enrollers entirely, or assigning them only to one group, introduces selection bias.<br>To address both problems we used a clone, censor, and weight approach, under which we created a dataset that duplicated (cloned) each eligible client at time zero representing a hypothetical randomisation and assigned one clone to each of the two dispensing strategies. Cloning ensures that clients assigned to a later enrolment window do not have to remain alive and in care through the full earlier window before entering the strategy, eliminating immortal time bias. However, cloning itself introduces informative censoring that must be addressed through weighting, as described below. |
| S5 | Cloning Procedure and Artificial Censoring | At time zero, each eligible client was duplicated into two identical copies. One copy was assigned to the early enrolment strategy (initiate 6MMD 0–3 months after time zero) and the other to the standard enrolment strategy (initiate 6MMD 3–9 months after time zero). Both copies carried identical baseline characteristics, as they were derived from the same client.<br>We then applied artificial censoring when a copy deviated from the dispensing strategy assigned at time zero. For instance, if a client enrolled in 6MMD 5 months after time zero, the copy assigned to the early enrolment strategy (0–3 months) was censored at 3 months, because at that point the client was no longer following that strategy. The copy assigned to the standard enrolment strategy (3–9 months) was not censored and was followed for its entire eligible follow-up period, as initiating at 5 months falls within the standard window.<br>A client who never initiates 6MMD but met the outcome definition at 5 months after time zero would be censored at 3 months for the early enrolment copy (because they did not initiate within the 0–3 month window), but would be recorded as having met the outcome at 5 months for the standard enrolment copy, because they were still eligible for the standard strategy at the time |
|  |  | of the event (5 months falls within the 3–9 month window). This censoring approach eliminates immortal time bias by ensuring no copy accumulates follow-up time under a strategy it is no longer following.<br>Supplementary figure 1 illustrates the follow-up schema for seven hypothetical clients, showing their original observations and the strategy copies they were assigned. |
| S6 | **Inverse Probability of Censoring Weights (IPCW)** | Artificial censoring is potentially informative: the type of client who deviates from their assigned strategy and is therefore censored may differ systematically from those who remain on-strategy. If left unaddressed, this informative censoring biases effect estimates. Stabilised inverse probability of censoring weights (IPCW) were estimated to correct for this.<br>Data were expanded from one row per clone into a person-month format, with one row per clone per month of follow-up. For each month, two pooled logistic regression models were fitted, with the binary outcome being whether the clone remained uncensored (i.e. did not deviate from the assigned strategy) in that month.<br>The **numerator model** included follow-up month and treatment strategy only, capturing the marginal (unadjusted) probability of remaining on-strategy at each time point. The **denominator model** additionally included age category at ART initiation, sex, size of facility ART cohort, and urban or rural facility location, capturing the conditional probability of remaining on-strategy given client and facility characteristics.<br>Stabilised IPCW were computed as the ratio of the predicted probability from the numerator model to the predicted probability from the denominator model in each month of follow-up. Stabilisation using a non-trivial numerator rather than 1, reduces variability in the weights compared with unstabilised weights, improving precision without introducing bias. Cumulative stabilised IPCW were obtained by taking the running product of these monthly ratios across all follow-up months for each clone, estimated separately within each strategy arm. |
| S7 | **Inverse Probability of Treatment Weights (IPTW)** | Inverse probability of treatment weights (IPTW) were estimated to address baseline confounding between clients who enrolled early and those who enrolled at standard timing. Baseline characteristics that predict both the timing of 6MMD enrolment and the risk of treatment interruption could bias comparisons if not accounted for.<br>The propensity score the conditional probability of early 6MMD enrolment given baseline covariates was estimated using logistic regression with the following covariates: age category at ART initiation, sex, facility ART cohort size, and urban or rural facility location.<br>Stabilised IPTW were calculated as the ratio of the marginal probability of the observed treatment to the conditional probability of the observed treatment given covariates. For early enrollers, the weight was calculated as the overall proportion of clients enrolled early divided by the propensity score. For standard enrollers, the weight was calculated as the overall proportion enrolled at standard timing divided by one minus the propensity score. Stabilisation using the marginal treatment probability in the numerator (rather than 1) reduces weight variability while preserving unbiasedness.<br>Covariate balance between strategy groups after IPTW weighting was assessed using standardised mean differences (SMD). An SMD of less than 0.10 for each covariate was considered indicative of adequate balance, suggesting that the weighted groups were comparable on measured baseline characteristics. |
| S8 | **Combined Weight and Trimming** | The final analytic weight for each clone at each month of follow-up was calculated as the product of the IPTW and IPCW (Final weight = $\text{IPTW} \times \text{IPCW}$). This combined weight simultaneously addresses baseline confounding through the IPTW component and informative censoring from protocol deviation through the IPCW component, producing estimates that approximate those that would be obtained from a randomised trial with full protocol adherence.<br>To limit the influence of extreme weight values, trimming was applied in three stages. First, IPTW were trimmed independently at the 99th percentile of the IPTW distribution. Second, IPCW were trimmed independently at the 99th percentile of the IPCW distribution. Third, the combined weight ($\text{IPTW} \times \text{IPCW}$) was trimmed at the 99th percentile of the combined weight distribution. Trimming of each component was applied prior to multiplication; the additional trimming of the combined weight addressed any residual extreme values arising from the product of the two components.<br>The distribution of the final analytic weights is summarised in Supplementary Table S1 |
| S9 | **Outcome Model: Pooled Logistic Regression** | The primary outcome model was a pooled logistic regression (PLR) fitted to a person-month dataset created by splitting each clone's follow-up into one-month intervals, producing one row per clone per month of follow-up. The binary outcome in each interval was whether treatment interruption occurred in that month.<br>The PLR included indicator variables for each follow-up month to account for the baseline discrete-time hazard the underlying probability of treatment interruption varying by time since time zero and treatment strategy (early versus standard enrolment) as the main exposure of interest. Under the rare-event assumption, the per-interval odds ratio from a PLR approximates the hazard ratio from a continuous-time proportional hazards model. This assumption was satisfied in our data: per-interval treatment interruption probabilities were well below 5% across all follow-up months. PLR was selected because it naturally accommodates the clone–censor–weight framework in discrete-time person-month data and permits direct estimation of standardised cumulative incidence and risk differences.<br>Robust sandwich standard errors were used throughout to account for within-client correlation introduced by cloning, as each eligible client contributes two copies to the analysis.<br>Two models were fitted. **Model 1** applied IPCW only, adjusting for informative censoring arising from protocol deviation during follow-up but not for baseline confounding between early and standard enrollers. **Model 2** applied the combined IPTW × IPCW weight, simultaneously adjusting for both informative censoring and baseline confounding. Comparison of estimates across models allowed assessment of the contribution of baseline confounding to the observed association: attenuation of the odds ratio in Model 2 relative to Model 1 was interpreted as evidence of baseline confounding, while stability of estimates across models was taken as evidence that the censoring-adjusted estimate was not materially biased by baseline differences between groups. |
| S10 | **Absolute Effect Measures** | Predicted cumulative incidence of treatment interruption at 18 months from time zero was estimated separately for each strategy arm from PLR Model 2, using the discrete-time survival product formula: $S(t) = \prod [1 - h(t)]$, where $h(t)$ is the predicted monthly probability of treatment interruption from the PLR at each time point $t$ and the product is taken from time zero through 18 months from time zero.<br>The risk difference was calculated as the difference in cumulative incidence between the early and standard enrolment strategies at 18 months from time zero: Risk difference = Cumulative incidence (early enrolment) – Cumulative incidence (standard enrolment). |
| S11 | **Sensitivity Analysis** | Weighted Poisson regression models with a log person-time offset were fitted as an internal consistency check, under both weighting schemes: Model 1 (IPCW only) and Model 2 (combined IPTW × IPCW). The Poisson models estimated incidence rate ratios (IRR) comparing early versus standard enrolment.<br>Under the rare-event assumption which was satisfied in our data the per-interval odds ratios from the PLR models and the IRRs from the Poisson models were expected to be close. Close agreement between the two model types provides an internal check on the robustness of the primary analysis and confirms that the choice of outcome model does not materially affect conclusions.<br>Sensitivity to confounding adjustment was additionally assessed by comparing Model 1 and Model 2 estimates across both the PLR and Poisson specifications. Attenuation of the odds ratio or IRR after IPTW adjustment was interpreted as evidence of residual baseline confounding in the censoring-adjusted estimate. Stability of estimates across models was taken as evidence that the IPCW-adjusted estimate was not materially biased by baseline differences between early and standard enrollers. |

**Supplementary table 2.** Covariate balance before and after IPTW weighting.

| Covariate | Unweighted SMD | Weighted SMD |
| --- | --- | --- |
| Age category | 0.036 | 0.005 |
| Sex | -0.103 | -0.007 |
| Facility ART patient volume (TROA) | -0.410 | -0.003 |
| Urban/rural status | 0.056 | 0.001 |

*SMD = Standardised Mean Difference. Values close to 0 indicate good balance between Early and Standard 6MMD groups. Unweighted SMD: difference before applying IPTW. Weighted SMD: difference after applying stabilised IPTW. IPTW = Inverse Probability of Treatment Weighting, derived from logistic regression of Early 6MMD enrolment on age category, sex, facility type (TROA), and urban/rural status. Threshold for acceptable balance: SMD < 0.10. All covariates achieved balance after weighting.*

**Supplementary table 1.** Weighting diagnostics and covariate balance assessment.

|  | Propensity Score |  | IPTW |  | IPCW |  | IPTW × IPCW |  |
| --- | --- | --- | --- | --- | --- | --- | --- | --- |
| Measure | Early<br>(3–<6m) | Standard<br>(6–12m) | Early<br>(3–<6m) | Standard<br>(6–12m) | Early<br>(3–<6m) | Standard<br>(6–12m) | Early<br>(3–<6m) | Standard<br>(6–12m) |
| Median | 0.328 | 0.265 | 0.970 | 0.928 | 1.005 | 1.007 | 0.933 | 0.934 |
| IQR | 0.27–0.44 | 0.24–0.38 | 0.72–1.20 | 0.90–1.10 | 1.00–1.01 | 1.00–1.01 | 0.88–1.20 | 0.89–1.19 |
| Mean (SD) | 0.349<br>(0.101) | 0.306<br>(0.092) | 0.992<br>(0.287) | 1.003<br>(0.153) | 0.995<br>(0.020) | 1.002<br>(0.008) | 0.994<br>(0.206) | 0.999<br>(0.202) |
| Min | 0.201 | 0.201 | 0.625 | 0.853 | 0.914 | 0.968 | 0.611 | 0.622 |
| Max | 0.606 | 0.509 | 1.418 | 1.388 | 1.005 | 1.007 | 1.425 | 1.427 |

*PS = Propensity score: predicted probability of Early 6MMD enrolment from logistic regression on age category, sex, facility type (TROA), and urban/rural status. N=2,318 with non-missing covariates (13 excluded). IPTW = Inverse Probability of Treatment Weighting (baseline confounding). Stabilised weights calculated at patient level (Early n=741, Standard n=1,577). Eight IPTW values were truncated at the 99th percentile (1.418). Stabilised IPTW ranged from 0.625 to 1.418 and showed limited variability (IQR 0.72–1.20 for early enrollers and 0.90–1.10 for standard enrollers).*

**Supplementary figure 1.**
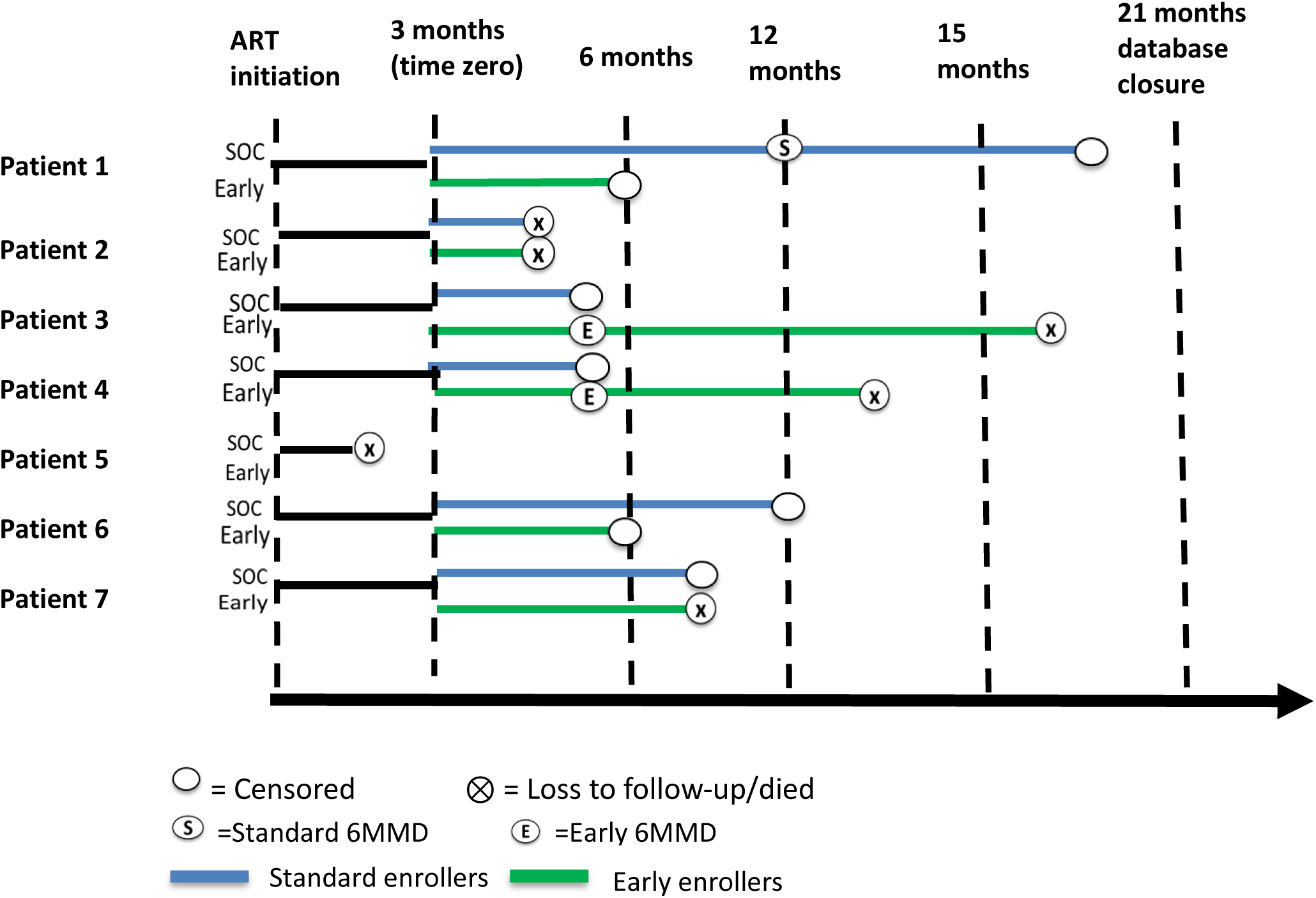
Follow-up schema for the clone and censor approach using seven hypothetical clients in a target trial emulation comparing early vs standard 6MMD initiation in Zambia

## Notes

### Competing Interest Statement

The authors have declared no competing interest.

### Summary of Updates

We revise because our funder Gates Foundation required CC BY license.

